# SWATH-MS based proteomic profiling of Pancreatic Ductal Adenocarcinoma tumours reveals the interplay between the extracellular matrix and related intracellular pathways

**DOI:** 10.1101/2020.06.04.20116640

**Authors:** EE Nweke, P Naicker, S Aron, S Stoychev, J Devar, DL Tabb, OJ Jones, MD Smith, GP Candy

## Abstract

Pancreatic cancer accounts for 2.8% of new cancer cases worldwide and is projected to become the second leading cause of cancer-related deaths by 2030. Patients of African ancestry appear to be at an increased risk for pancreatic ductal adenocarcinoma (PDAC), with worse severity and outcomes. The purpose of this study was to map the proteomic and genomic landscape of a cohort of PDAC patients of African ancestry.

Thirty tissues (15 tumours and 15 normal adjacent tissues) were obtained from consenting South African PDAC patients. Optimisation of the sample preparation method allowed for the simultaneous extraction of high-purity protein and DNA for SWATH-MS and OncoArray SNV analyses.

We quantified 3402 proteins with 49 upregulated and 35 downregulated proteins at a minimum 2.1 fold change and FDR adjusted p-value (q-value) ≤ 0.01 when comparing tumour to normal adjacent tissue. Many of the upregulated proteins in the tumour samples are involved in extracellular matrix formation (ECM) and related intracellular pathways. Proteins such as EMIL1, KBTB2, and ZCCHV involved in the regulation of ECM proteins were observed to be dysregulated in pancreatic tumours. Approximately 11% of the dysregulated proteins, including ISLR, BP1, PTK7 and OLFL3, were predicted to be secretory proteins. Additionally, we identified missense mutations in some upregulated proteins, such as MYPN, ESTY2 and SERPINB8. These findings help in further elucidating the biology of PDAC and may aid in identifying future plausible markers for the disease.

## Introduction

Pancreatic ductal adenocarcinoma (PDAC) originates from the ductal epithelial cells of the pancreas and accounts for 85% of all pancreatic cancers. Worldwide, the incidence and mortality rates of PDAC are rising compared to other cancers (1). Despite current therapeutic strategies, the survival rate is dismal, with a five-year survival rate of 8% (2). These poor statistics are chiefly due to late detection, allowing for invasion and metastasis of cancer, and therapeutic resistance. In multiracial countries such as the United States of America, African Americans have increased incidence and poorer survival rates compared to other ethnicities. Although this has been largely attributed to social factors such as smoking, alcohol consumption, obesity and diabetes mellitus, current studies have determined genetics as an underlying factor (3–5).

The tumour microenvironment (TME) of PDAC plays a role in enabling chemotherapeutic resistance, cancer cell proliferation, invasion, migration and metastasis. It is characterised by a dense extracellular matrix (ECM) structure, stromal cells, cancer-associated fibroblasts and immune cells (6–10). The overexpression of ECM proteins exacerbates PDAC tumourigenesis, fostering tumour growth. Pancreatic cancer cells secreting ECM proteins such as collagen and fibronectin, were found to have increased proliferation and were desensitized to chemotherapeutic drugs such as gemcitabine (11–14). Hence, there is a need for the identification of novel proteins involved in the ECM formation that could be potential chemotherapeutic targets and biomarkers of PDAC.

Mass spectrometry-based technologies have proven useful in the identification of proteins with important roles in PDAC progression (15). Sequential window acquisition of all theoretical fragment ion spectra (SWATH) is a label-free application of mass spectrometry (MS), suitable for discovery proteomics where hundreds or thousands of proteins are quantified at high analyte throughput and reproducibility (Gillet et al, 2012, Ludwig et al., 2018).

In the present study, protein and DNA were simultaneously extracted from the same tissue samples, which allowed SWATH-MS and OncoArray single nucleotide variation (SNV) profiling of tissues obtained from resectable (stage I and II) South African PDAC patients who underwent a Whipple procedure. Several significantly differentiated proteins were identified, many of which were enriched in the extracellular matrix formation and related intracellular pathways. By querying databases, some of the identified proteins were shown to be secreted into the bloodstream which could make them potential biomarkers of PDAC.

## Methods

### Ethics statement

The Human Research Ethics Committee of the University of Witwatersrand approved this study (HREC-M150778). Each patient gave written consent for sample collection and recording of demographic and clinical data.

### Patient recruitment and sample collection

Biopsy samples were obtained from 15 consenting South African patients (aged between 53-95 years old) with histologically confirmed PDAC at the Chris Hani Baragwanath Academic Hospital (Johannesburg, South Africa). All biopsies were obtained immediately after Whipple procedures to resect of the pancreas. One core tumour biopsy and one adjacent ‘normal’ tissue (> 2cm away from the tumour) were obtained (30 samples) and immediately frozen at −80°C (**Table S1**). The overview of the study workflow is shown in **Figure 1**.

**Figure 1:**
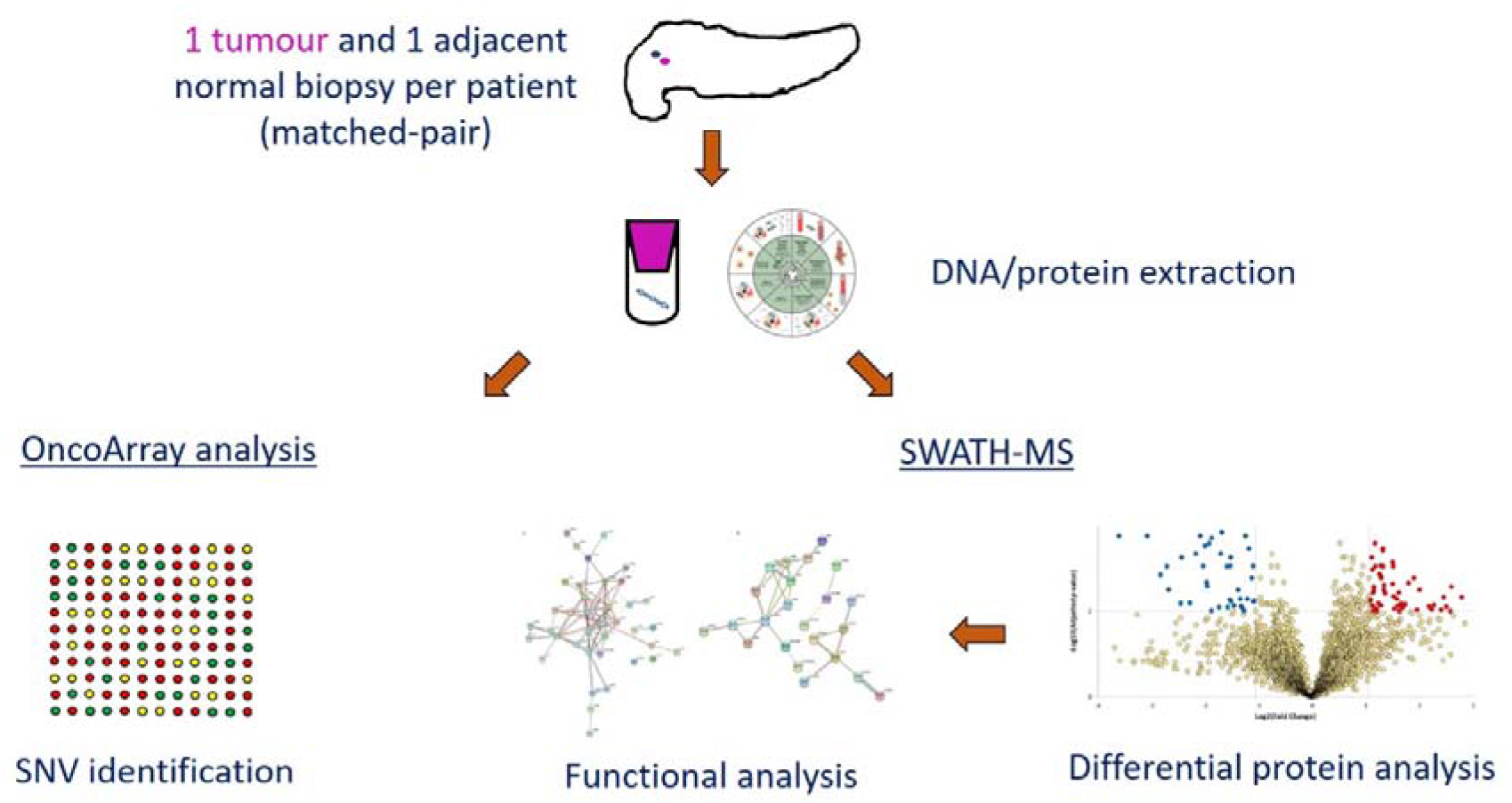
Overview of study workflow, from sample collection to functional analysis.

### Tissue processing

Tissues (15-35mg) were homogenised in 600μl of ATL lysis buffer (Qiagen Hilden, Germany) supplemented with 40mM DTT, using a Tissue Ruptor (Qiagen). The homogenised solution was centrifuged at 14 000*g* for 3 mins to remove any micro-tissue particulate from the solution. From the recovered lysate 180μl was taken for DNA purification.

### Protein preparation for mass spectrometry

The remainder of the lysate was incubated with four parts pre-chilled acetone for 30 min at −20°C. The mixture was centrifuged at 14 000g at 4°C for 10 min and the resultant pellet was washed with ice-cold ethanol. The pellet was resuspended in 50mM Tris-HCl pH 8.0 containing 4% sodium dodecyl sulphate (SDS) and a Roche complete™ EDTA-free protease inhibitor cocktail. The protein concentration was measured using the Pierce Bicinchoninic assay (Thermo Fisher Scientific, Massachusetts, USA). Aliquots of protein solution were then stored at −80°C until further processing.

Protein samples were thawed and 20μg per sample was reduced with 10mM dithiothreitol (VWR Pennsylvania, United States) at 37°C for 30 min and alkylated with 40mM iodoacetamide at room temperature in the dark for 30 min. The samples were then purified of detergents and salts using MagReSyn™ HILIC microparticles (ReSyn Biosciences Edenvale, South Africa) in a 96 deep-well plate for automated sample preparation of 12 samples in parallel using the KingFisher Duo™ system (Thermo Fisher Scientific), as previously described (17). Briefly, magnetic hydrophilic affinity microparticles (20μl, 200μg) were equilibrated in 200μl of 100mM ammonium acetate pH 4.5 containing 15% acetonitrile (MeCN). Microparticles were then transferred to 100μl of protein binding solution (each protein sample adjusted to 50μl using 4% SDS and added to 50ul of 200mM ammonium acetate pH 4.5 containing 30% MeCN) and mixed for 30 min at room temperature. The captured proteins were washed twice in 200μl of 95% MeCN and transferred to 200μl of 25mM ammonium bicarbonate containing 2μg of sequencing grade trypsin (Promega, Madison, USA) and mixed for 4 hrs digestion at 37°C. The automated on-bead protein capture, clean-up and digest protocol was programmed using BindIt software v4.0 (Thermo Fisher Scientific) and is available upon request and further described in **Figure S1**. Resultant peptides were dried and stored at −80°C before Liquid Chromatography-MS analysis.

### Liquid chromatography-mass spectrometry (LC-MS) data acquisition

Approximately 1ug of tryptic peptides per sample was analysed using a Dionex Ultimate 3000 RSLC system coupled to an AB Sciex 6600 TripleTOF mass spectrometer (AB Sciex, Massachusetts, USA). For data-dependent acquisition (DDA, used for spectral library building), tumour and normal samples were each pooled in their respective groups and spiked with Biognosys iRT retention time peptide standards. Four injection replicates of each group were acquired (*n* = 8). For SWATH, each sample was injected once. Peptide samples were inline de-salted using an Acclaim PepMap C18 trap column (75μm × 2cm; 2 min at 5μl.min^-1^ using 2% Acetonitrile, ACN/0.2% FA). Trapped peptides were gradient eluted and separated on an Acclaim PepMap C18 RSLC column (75μm × 15cm, 2μm particle size) at a flow-rate of 0.5μl.min^-1^ with a gradient of 4-40% B over 60 min (A: 0.1% FA; B: 80% ACN/0.1% FA). For DDA, precursor (MS) scans were acquired from *m/z* 400-1500 (2^+^-5^+^ charge states) using an accumulation time of 250ms followed by 80 fragment ion (MS/MS) scans, acquired from *m/z* 100-1800 with 25ms accumulation time each. For SWATH, precursor scans ranged from *m/z* 400 to 900 using 100 variable-width windows that overlapped by 0.5 Da, and fragment ions were acquired from *m/z* 100-1800 with 25ms accumulation time per window.

### Liquid Chromatography-Mass Spectrometry (LC-MS) Data analysis

Raw DDA files (.wiff), *n* = 8, were included in a combined search using Protein Pilot software v5.0.2.0 (AB Sciex, Massachusetts, USA), against a database of human reference proteome contained in SwissProt (Accessed 9 June 2019) supplemented with sequences of common contaminants and iRT peptide retention time standards (Biognosys Schlieren, Switzerland). This searched data file was imported into Skyline software v19.1 (Maclean et al., 2010) with peptide features extracted to build a spectral library. A cut-off score of 0.994 (from Protein Pilot report) corresponding to 1% local peptide false discovery rate (FDR) was applied for a library building, with fixed carbamidomethylation of cysteine and variable N-terminal acetylation being the only allowable modifications. The minimum peptide length was 7 and the maximum 36 amino acids and enzyme for digestion set as trypsin with one allowable missed cleavage. The built spectral library contained 141 775 transitions matching to 19 861 peptides and 3413 proteins. Decoy entries were generated by shuffling the peptide sequence, with 141 775 decoy transitions and 19 861 decoy peptides added.

SWATH data files (.wiff), *n* = 30, were centroided and converted to mzML format using the msconvert tool in the ProteoWizard software suite v3.0.19217 (18). The converted data files were imported into Skyline and matched to the spectral library for correct assignment of the mass spectra. A fragment ion mass tolerance of 0.5m/z was employed for library matching, with a minimum of three and maximum of six of the most intense product ions being selected. The b and y ions with +1 and +2 charge states matching to peptides with +2, +3 and +4 charge states were allowed and product ions from b3 to the last ion were allowed for selection. The iRT peptides were used to align retention times across the dataset using the score to run a regression. A 2 min window around the predicted retention time was used to assist in identification. A peak scoring model was trained using mProphet (19) and the decoy peptides generated from the spectral library. All peaks were re-integrated using this model. The MSstats (20) group comparison plugin was implemented within Skyline to perform quantitative analysis. A normalisation method (equalise medians) was applied for the comparisons across the experimental groups. The MSstats design sample size plugin was used to determine the statistically significant fold change that could be observed based on the desired power, sample number and variability of the dataset. The filtered list of dysregulated proteins was manually inspected within Skyline to verify the quality of the precursors and transitions used for quantitation. Principal component analysis (PCA) was performed using the peptide-level quantification of each sample within Perseus v1.6.5.0 (21).

### DNA extraction and SNV array

DNA purification was performed as per the manufacturer’s instruction using the Qiagen DNeasy Blood and Tissue kit (Qiagen Hilden, Germany). The total DNA was quantified using a NanoDrop ND-1000 UV spectrophotometer. A ratio of absorbance at 260nm:280nm of >1.9 and 260nm:230nm of >1.5 was observed in all samples. The Illumina Infinium® 0ncoArray-500k Beadchip (Ilumina California, United States) was used for single nucleotide variation (SNV) analysis using 200ng of DNA for each sample. The SNV array was conducted as per manufacturer’s instruction. GenomeStudio (https://emea.illumina.com/techniques/microarrays/) was used to perform clustering and QC of the raw intensity data and the genotype calls were exported as a GenomeStudio report. For downstream analysis, the GenomeStudio report was converted into PLINK (22) format using the *topbottom* module of the H3ABioNet workflow (https://github.com/h3abionet/h3agwas). The original dataset consisted of 499 170 SNVs genotyped for 48 samples. Some samples were genotyped in replicate and the replicates were utilised to improve the genotype calling when converting the data to PLINK format.

### Single nucleotide variation (SNV) analysis

The genotype data were subset to include only tumour-normal pairs and replicates were removed, as related samples cannot be used in the association tests. Quality control was conducted on the dataset to remove low-quality SNVs and samples. SNVs with greater than 1% missingness, lower than 1% minor allele frequency and deviations from Hardy-Weinberg Equilibrium were removed. Tumour samples were coded as cases and normal samples coded as controls.

### Functional analysis

PANTHER v14.1 (23) and REACTOME v70 (24) were applied to show pathway-enrichment analysis of identified dysregulated proteins. Cytoscape v3.8.0 (25) with the STRING (26) plug-in was used to show network interaction between dysregulated proteins. ShinyGO v0.61 (27) was used for graphical representation of enriched biological processes/pathways and the Ensembl Variant Effect Predictor tool (28) modelled the consequences of observed SNPs.

## Results

### Differential protein analysis in tumours

Using SWATH-MS, we matched 3413 proteins and 19 861 peptides to our in-house generated spectral library. Peptide quantification was performed on MS2 ion measurements using Skyline (Maclean et al., 2010). A principal component analysis (PCA) determined the maximum covariance among the samples based on the abundance of all peptides measured in the analysis. This allowed for unsupervised grouping of samples without any manual assignment of groups. The PCA plot (**Figure S2**) shows two defined groups of samples in agreement with the two clinically defined groups in the study. The normal adjacent group dataset shows wider distribution compared to the tumour group dataset.

An MSstats group comparison was performed with Skyline software to determine the observed differential abundance (fold change) of each quantifiable protein across the tumour and normal adjacent experimental groups. A minimum fold change ≥2.1 and maximum FDR adjusted p-value (q-value) ≤0.01 was used to filter proteins that were significantly different between tumour and corresponding normal tissues. The fold change threshold was calculated using the MSstats design sample size plugin within Skyline, based on 0.8 power, 0.01 FDR and 15 replicates per group (**Figure S3**). We found 49 upregulated and 35 downregulated proteins in tumours compared to normal adjacent tissues (**Figure 2, Table S2**). Across the complete dataset of quantified proteins (3402), 318 are predicted to be secreted based on information from the Human Protein Altas (29) and Uniprot (30), both assessed on March 24^th^ 2020. Among the dysregulated proteins, 78% of the observed differentially expressed proteins (DEPs) have been shown to have prognostic potential in various cancers like colorectal, renal, liver, lung, urothelial, thyroid and endometrial. AP2A2, PLST, PLSI, BLVRB and SLC2A1 were known intracellular prognostic markers of pancreatic cancer (**Table S2**). Additionally, about 11% were predicted to be secreted into the bloodstream, making them potential reporters for tumour progression and metastasis. These include proteins such as ISLR, OLFL3, PTK7, GBB1, HEM2, BP1, RL39, CAMP, PF4V, and LTBP1, which were reported as prognostic pancreatic cancer biomarkers in the Human Protein Atlas database.

**Figure 2:**
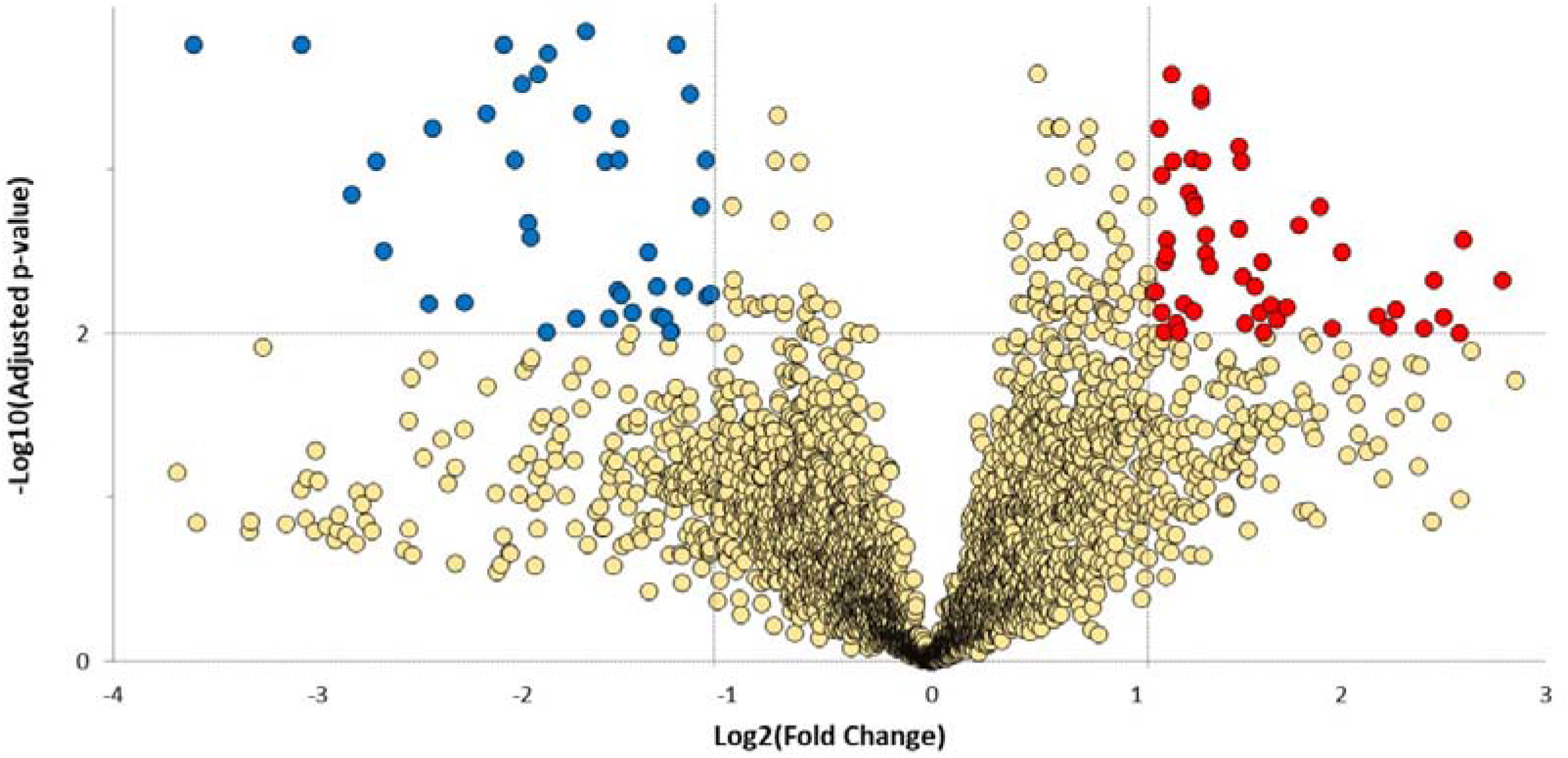
Volcano plot showing dysregulated proteins. Red and green nodes indicate downregulated and upregulated proteins group (based on a minimum fold change ≥2.1 and a maximum adjusted p-value (q-value) ≤0.01) in tumour compared to normal adjacent, respectively.

### Functional and network analysis of differentially expressed proteins

We observed that dysregulated proteins (**Table S2**) share several key biological processes (**Figure 3**) and pathways (**Table 1, 2**). These were mostly linked to the extracellular matrix formation/organisation and related intracellular signalling pathways (**Figure 4**). Top pathways enriched for the upregulated proteins included recycling pathway of L1, cell-extracellular matrix interactions and cell junction organisation. Top downregulated pathways were involved in the take-up of oxygen and the release of carbon dioxide from erythrocytes.

**Figure 3:**
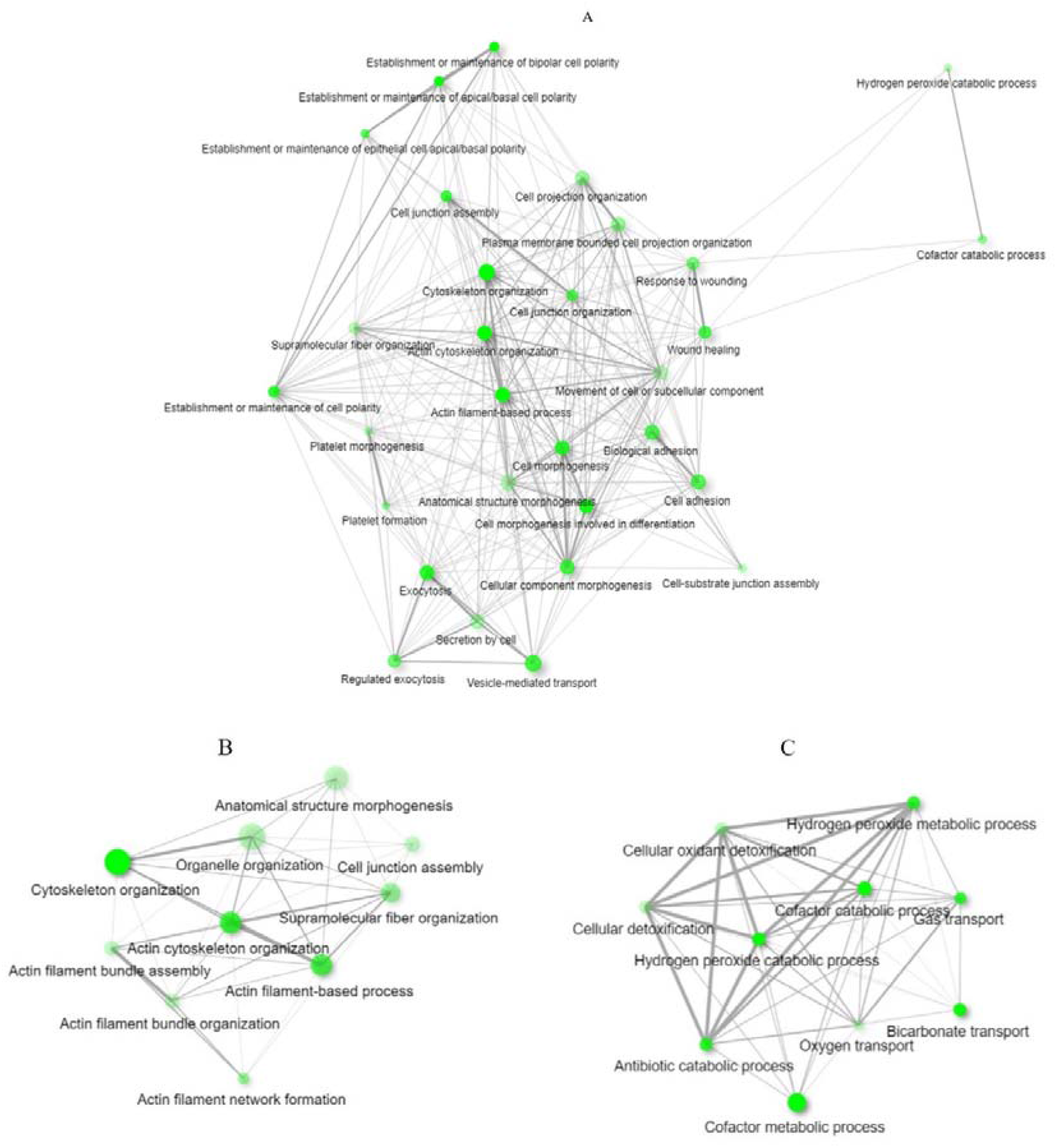
The interaction network analysis of the relationship between biological processes that are enriched by (a) dysregulated proteins. Separate analyses of (b) upregulated and (c) downregulated proteins are also shown. An edge indicates that two processes share 20% or more proteins. Thicker edges (lines) show that there are more overlapped edges. Bigger and darker nodes represent larger protein sets and more significantly enriched proteins, respectively. The plot was generated from ShinyGO.

**Table 1:** Top 10 significantly upregulated pathways in tumour samples. Generated from Reactome.

| Pathway name | False discovery rate | Submitted proteins |
| --- | --- | --- |
| Recycling pathway of L1 | 2.57E-05 | AP2A2, TBB6, TBB2A, TBB8, ACTG, DPYL2 |
| Cell-extracellular matrix interactions | 2.02E-04 | ACTG, ACTN1, LIMS1, ILK |
| RHO GTPases activate IQGAPs | 9.95E-04 | TBB6, TBB2A, TBB8, ACTG |
| Translocation of SLC2A4 (GLUT4) to the plasma membrane | 9.95E-04 | TBB6, TBB2A, TBB8 |
| MHC class II antigen presentation | 1.86E-03 | AP2A2, TBB6, TBB2A, CATB, CATC |
| Cell junction organisation | 2.30E-03 | PLEC, ACTG, ACTN1, LIMS1, ILK |
| Hemostasis | 5.73 E-03 | WDR1, TBB6, TBB2A, SPB8, TBB8, GBB2, GBB1, CAP1, ACTN1, ISLR, AT2B4 |
| Cell-Cell communication | 5.73 E-03 | PLEC, ACTG, ACTN1, LIMS1, ILK |
| Aggrephagy | 1.21E-02 | TBB6, TBB2A, TBB8 |
| Platelet activation, signalling and aggregation | 1.21E-02 | WDR1, ACTN1, ISLR, GBB2, GBB1 |

**Table 2:** Significantly downregulated pathways in tumour samples. Generated from Reactome.

| Pathway name | False discovery rate | Submitted proteins |
| --- | --- | --- |
| Erythrocytes take up oxygen and release carbon dioxide | 9.99E-09 | CAH1, HBB, HBA, B3AT, CAH2 |
| Erythrocytes take up carbon dioxide and release oxygen | 2.51E-08 | CAH1, HBB, HBA, B3AT, CAH2 |
| O2/CO2 exchange in erythrocytes | 2.51E-08 | CAH1, HBB, HBA, B3AT, CAH2 |
| Metabolism of porphyrins | 4.17E-03 | HEM2, BLVRB, HEM3 |
| Reversible hydration of carbon dioxide | 2.33E-02 | CAH1, CAH2 |
| Heme biosynthesis | 3.03E-02 | HEM2, HEM3 |

**Figure 4:**
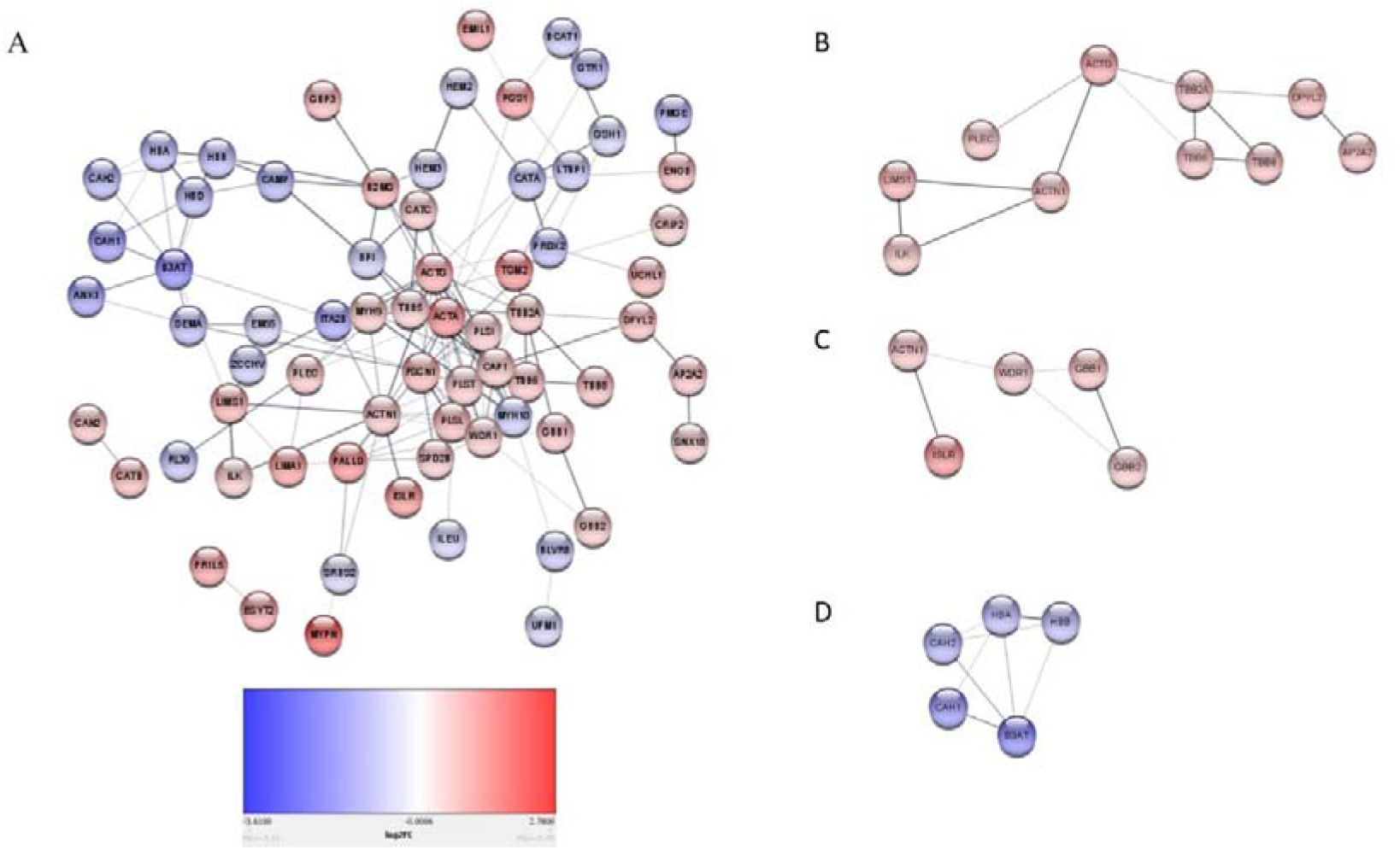
(A) The interaction network of the different dysregulated proteins. Proteins involved in Extracellular matrix formation/organisation (B) (Recycling pathway of L1, cell-extracellular matrix interactions, cell junction organisation, cell-cell communication, Platelet activation(C), and O_2_/CO_2_ transport(D) are also shown.

### Single nucleotide variants observed in tumours

The quality-controlled dataset contained 369, 259 SNVs for the 15 paired samples. Although the study was underpowered for detecting statistically significant associations due to the small sample size, we performed a simple linear regression association test on the complete dataset (all tumour versus normal adjacent) to identify SNVs with a difference in allele frequency between the tumour and normal adjacent groups. Top associated results are listed with p-values, although none of these was significant after correcting for multiple testing (**Table S3**).

To assess the possible role of SNVs in the regulation of protein levels, the association analysis was re-run on a subset of SNVs extracted based on the gene regions for the previously identified DEPs. The association results were assessed to identify SNVs with a difference in allele frequency between cases (tumour) and controls (normal adjacent). Extraction of SNVs in the gene regions identified from the proteomics results (significantly up and down-regulated proteins) resulted in 912 SNVs. A linear regression association test with this subset of SNVs was conducted and, although no significant difference in allele frequency was observed between the tumour and normal adjacent groups, SNVs were found within some DEPs (**Table S3**). We further annotated the types of SNVs (**Figure S4**). Sixty-nine percent of the coding SNVs found in DEPs were synonymous variants, while 31% were missense variants. Missense variants were found in *MYPN*, *SERPINB8* (SPB8) and *ESYT2*, which were among the top ten most upregulated proteins in the tumour group.

## Discussion

In this study, we performed proteomic and genomic profiling and identified proteins dysregulated in resected pancreatic tumours obtained from patients of African ancestry. For the first time, to our knowledge, dysregulation of proteins such as EMIL1, KBTB2, and ZCCHV, that are responsible for the regulation of ECM components, were observed in pancreatic tumours. Additionally, the secretion of some differentiated proteins into the bloodstream allows for them to be validated in future studies as potential biomarkers. Pathway analysis further showed that the majority of the DEPs were enriched in pathways responsible for, and related to extracellular matrix formation (**Figure 2**).

### Dysregulated proteins implicated in extracellular matrix formation

In this study, we observed that pathways involved in ECM formation and interactions were dysregulated corroborating the findings of several studies (31–34). The ECM modulates intracellular signalling pathways; consequently, aberrant ECM homeostasis and ECM remodelling can induce and enhance tumorigenesis. Furthermore, there is evidence that the altered expression of ECM components can affect intracellular signalling (35,36).

The enrichment of ECM-related pathways (Recycling pathway of L1, cell-extracellular matrix interactions, cell junction organisation, cell-cell communication) was due to the upregulation of specific proteins, such as PLEC, ACTN1, LIMS1, and ILK (**Table 1, Figure 4(B)**). We recently showed that these proteins were also overexpressed at the mRNA level (37) with increased activity of the MAPK and P13K/AKT signalling pathways (37). Both the MAPK and P13K/AKT signalling pathways are regulated by FAK/SRC activation which results from the interaction of integrin with ECM molecules (38). Drawing from our studies and literature, **Figure 5** shows a schematic of how these pathways interplay in the ECM.

**Figure 5:**
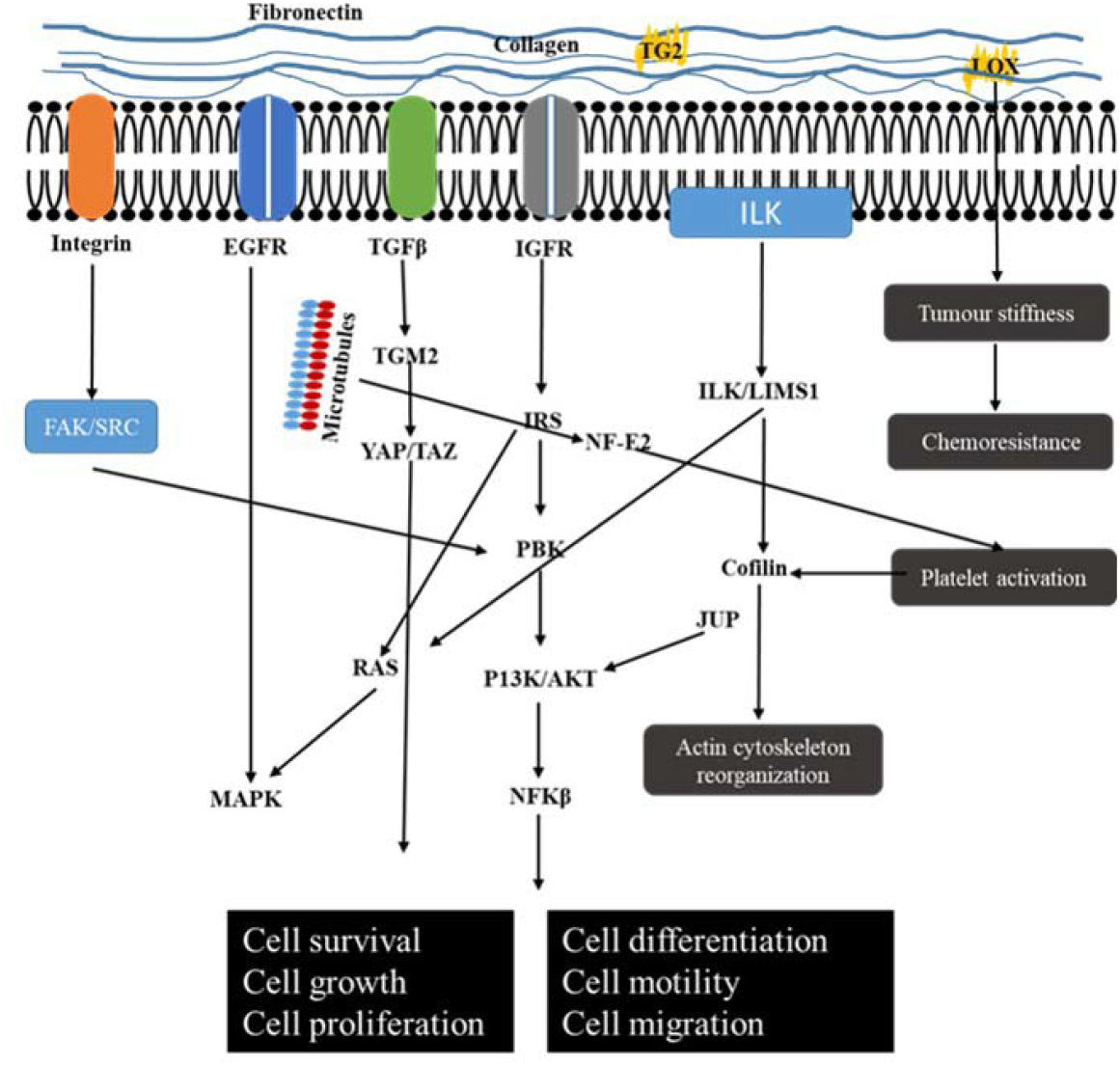
The schematic interplay between the extracellular matrix and intracellular signalling pathways.

The identified dysregulated proteins play important roles in tumorigenesis. PLEC (Plectin) was further found to be up-regulated in squamous cell carcinoma, aiding in the induction of malignant transformation from sinonasal inverted papilloma (39) and has been utilised in pancreatic cancer detection using targeted nanoparticles (40–42). High expression of ACTN1(Actinin Alpha 1) promotes cellular migration, invasion and metastasis, by weakening E-cadherin adhesion that ensures cellular integrity (43). Additionally, ACTN1 has been identified as a key regulator of PDAC progression, using a multidimensional systems-level analysis approach (44). Increased LIMS1 (LIM and senescent cell antigen-like-containing domain protein 1), a focal adhesion protein, has been associated with poor prognosis in laryngeal and pancreatic cancers (45,46). Also, during the knockdown of LIMS1, apoptosis was induced in breast cancer cells (47). In PDAC, it was found to promote cell survival under hypoxic conditions by activating the AKT/mTOR signalling pathway (45). ILK (Integrin-linked kinase) which interacts with LIMS1 (48) was also overexpressed in PDAC tumours. One study using neuroblastoma cell lines found that silencing the LIMS1/ILK pathway reduced cellular proliferation, highlighting its potential as a therapeutic target (49).

We also observed the dysregulation of several other proteins, such as TGM2, EMIL1, KBTB2, and ZCCHV, related to the ECM but not enriched in the top significantly dysregulated pathways. Compared to normal adjacent tissues, tumours showed a 6.04-fold increase in Transglutaminase (TGM2) expression, an enzyme involved in extracellular matrix stiffness, by cross-linking collagen 1 fibres (50,51). In PDAC tissues, transforming growth factor-beta induces TGM2 expression resulting in the activation of the YAP/TAZ signalling pathway, promoting cellular proliferation, invasion and metastasis (51,52). EMIL1 is an extracellular matrix glycoprotein that has been identified in both primary and metastatic colon tumours (53). KBTB2 (Kelch repeat and BTB domain-containing protein 2) is part of the kelch-repeat superfamily that functions in mediating the ubiquitination and degradation of target proteins. Importantly, members of this family are known to mediate cellular adhesion (54,55). One member of the family, keap1, was shown to target ectoplasmic specializations indicating interactions with ECM components (56). ZCCHV (or ZAP) functions in inhibiting viral replication by preventing viral mRNA accumulation (57,58). The anti-viral function of ZAP is regulated by Matrin 3, a member of the matrix metalloproteins known to be involved in the degradation of ECM components during tissue and embryo development (59,60). This study also found missense mutations in several upregulated proteins, including ESTY2. ESTY2 plays a role in cytoskeleton organisation, which may impact ECM and potentially the overall biology of PDAC cells. In lung cancer cells (61), the short and long variants of ESYT2 were implicated in the cortical distribution of actin and endocytosis, respectively.

Furthermore, we observed that upregulation of platelet activation, signalling and aggregation in tumours (Table 1, Figure 4(C)). Platelet activation results in the release of pro-angiogenic factors (such as VEGF and FGF) and pro-inflammatory markers (such as IL-8 and members of the C–X–C motif ligand family), inducing angiogenesis and inflammation, respectively (62–64). Of interest, pro-inflammatory factors contribute to the strengthening of the extracellular matrix in the tumour microenvironment (65). One study showed that elevated mean platelet volume (MPV) correlated with poor survival in pancreatic cancer patients (66). A major characteristic of platelet activation is the reorganisation of the actin cytoskeleton by cofilin-1 induction (67). Importantly, WDR1, which was overexpressed in this study, enhances the function of cofilin-1 (68,69).

In the present study, the downregulation of pathways involved in oxygen transport indicative of hypoxia (**Table 2, Figure 4(D)**) was observed. Hypoxia has been linked to tumour stroma stiffness that is characteristic of the extracellular matrix of pancreatic tumours. Tumour stiffness induces epithelial to mesenchymal transition, promoting chemo-resistance. This stiffness is primarily achieved when collagen fibres are cross-linked by lysyl oxidase (70), which is increased during hypoxia (Cox et al., 2013; Miller et al., 2015). Additionally, hypoxia is associated with poor survival in pancreatic cancer patients, promoting invasion and metastasis, even in the early stages of the disease, and enabling therapeutic resistance (74).

### Upregulation of proteins involved in cell division and metastasis

Several β-tubulin subtypes including TBB2A, TBB6 and TBB8 were significantly overexpressed in tumour samples (**Table 1)**. β-tubulin combine with α-tubulin to form microtubules, which play crucial roles in mitotic cell division (75) and cell adhesion (76). Drugs binding β-tubulin, such as colchicine and paclitaxel, are known to kill cancer cells (77). Increased expression and mutations in β-tubulin subtypes exacerbate drug resistance in cancer (78-81). Additionally, β-tubulin is expressed in platelets, aiding in their formation via the NF-E2 pathway (82).

## Conclusion

We have shown differentially expressed proteins in resected pancreatic ductal adenocarcinoma tumours obtained from patients of African ancestry using SWATH-MS. Many of these proteins are involved in cell proliferation and in the extracellular matrix formation/organisation, which plays a role in tumour progression and chemo-resistance. Targeting these proteins could be of effective therapeutic potential. Going forward, we plan to expand on the discovery dataset described here and further investigate the DEPs in a larger sample cohort. This would include analyses of biopsies at a single-cell level and plasma proteomics. In particular, the detection of secretory proteins in the bloodstream of PDAC patients may be beneficial in prognosis of the disease.

## Data Availability

The data files described in this study have been deposited to the ProteomeXchange Consortium via the PRIDE partner repository with the dataset identifier (PXD019549).

## Data availability

The data files described in this study have been deposited to the ProteomeXchange Consortium via the PRIDE partner repository (83) with the dataset identifier (PXD019549).

## Supplementary figures

**Figure S1:**
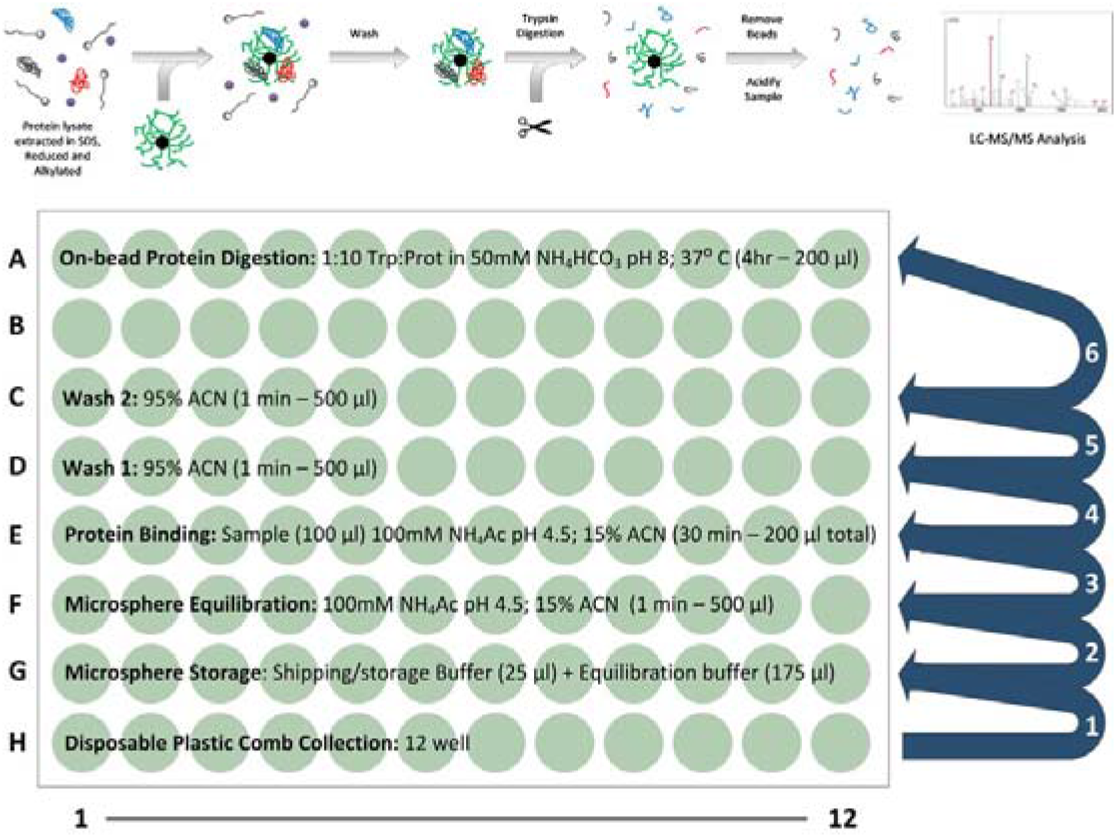
On-bead based protein capture, clean-up and digestion. Top: proteins are captured on magnetic hydrophilic affinity microparticles, followed by a high-organic wash to remove contaminants and on-bead digestion by addition of sequencing grade trypsin. Bottom: Plate set-up for automated sample processing in a Thermo Fisher Scientific KingFisher™ Duo Magnetic handling station.

**Figure S2:**
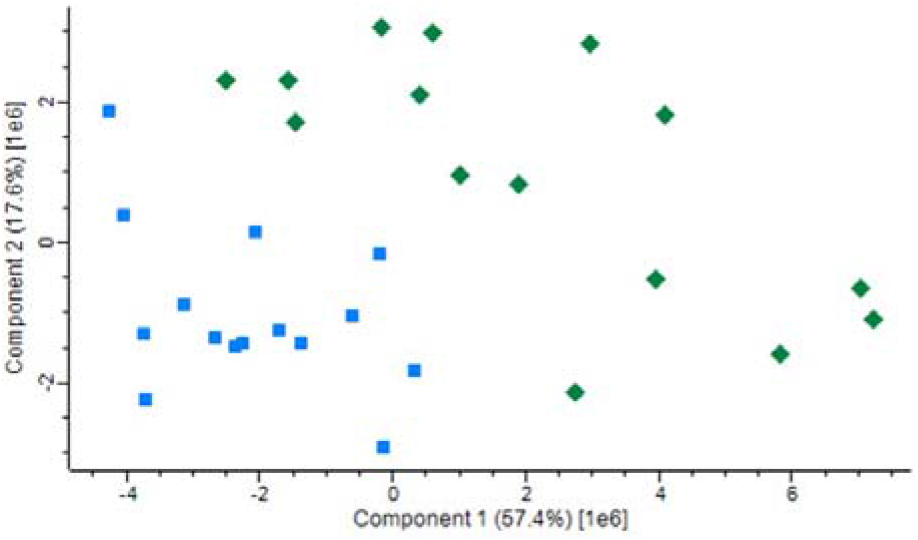
Principal component analysis (PCA) showing tumour samples as blue squares and normal adjacent samples as green diamonds. The PCA plot was generated using peptide abundance data of all peptides analysed per sample.

**Figure S3:**
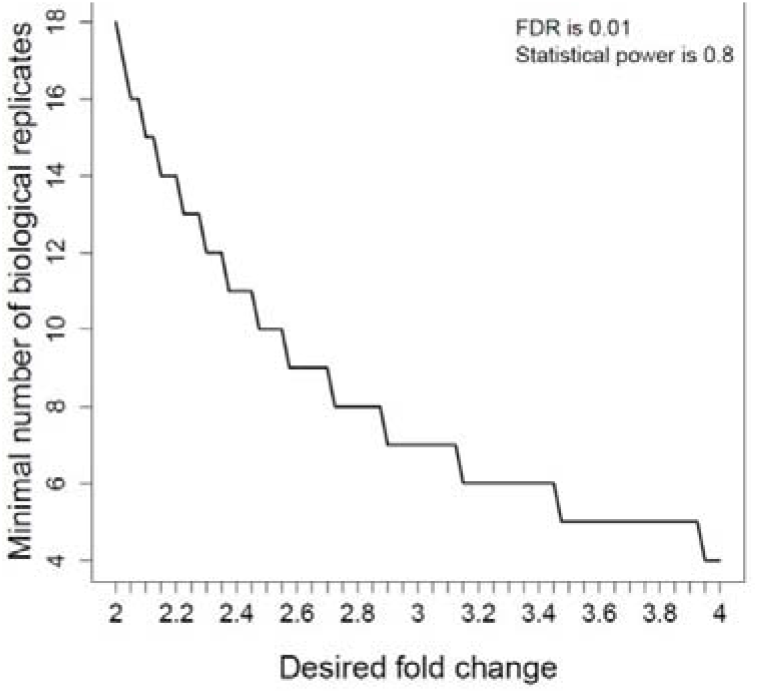
Sample size plot showing the relationship between the number of biological replicates and protein level fold change that can be distinguished based on the current dataset. A 2.1 fold change can be distinguished in the current study (biological replicates = 15) with statistical power set to 0.8 and false discovery rate (FDR) set to 0.01.

**Figure S4:**
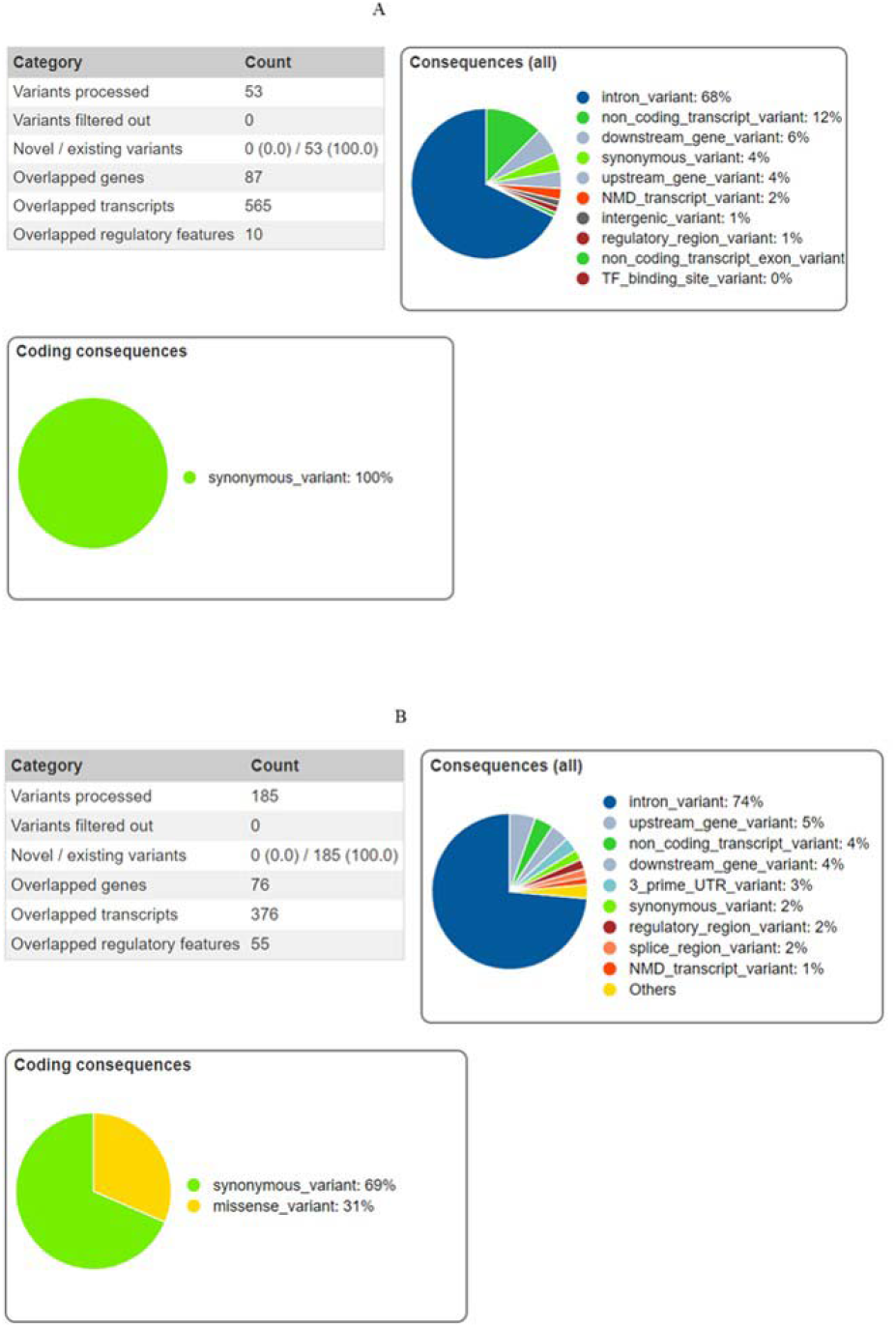
Types of variants observed from the list of SNVs found in (A) All Tumour vs Normal adjacent and (B) Tumour vs Normal adjacent in differentially expressed proteins only

## Supplementary tables

**Table S1:** Characteristics of patients recruited for the study

| Patient | Gender | Staging | TNM |
| --- | --- | --- | --- |
| 1 | Female | IIA | T2N1M0 |
| 2 | Male | IIA | T2N1M0 |
| 3 | Male | IIA | T2N1M0 |
| 4 | Male | IA | T1N0N0 |
| 5 | Female | IIB | T3N1M0 |
| 6 | Male | IIA | T2N1M0 |
| 7 | Male | IIA | T2N1M0 |
| 8 | Male | IIB | T3N1M0 |
| 9 | Female | IA | T1N0N0 |
| 10 | Male | IIA | T2N1M0 |
| 11 | Male | IIB | T3N1M0 |
| 12 | Female | IIA | T2N1M0 |
| 13 | Female | IIB | T3N1M0 |
| 14 | Female | IIB | T3N1M0 |
| 15 | Female | IIB | T3N1M0 |

TNM: TNM Classification of Malignant Tumors

**Table S2:**
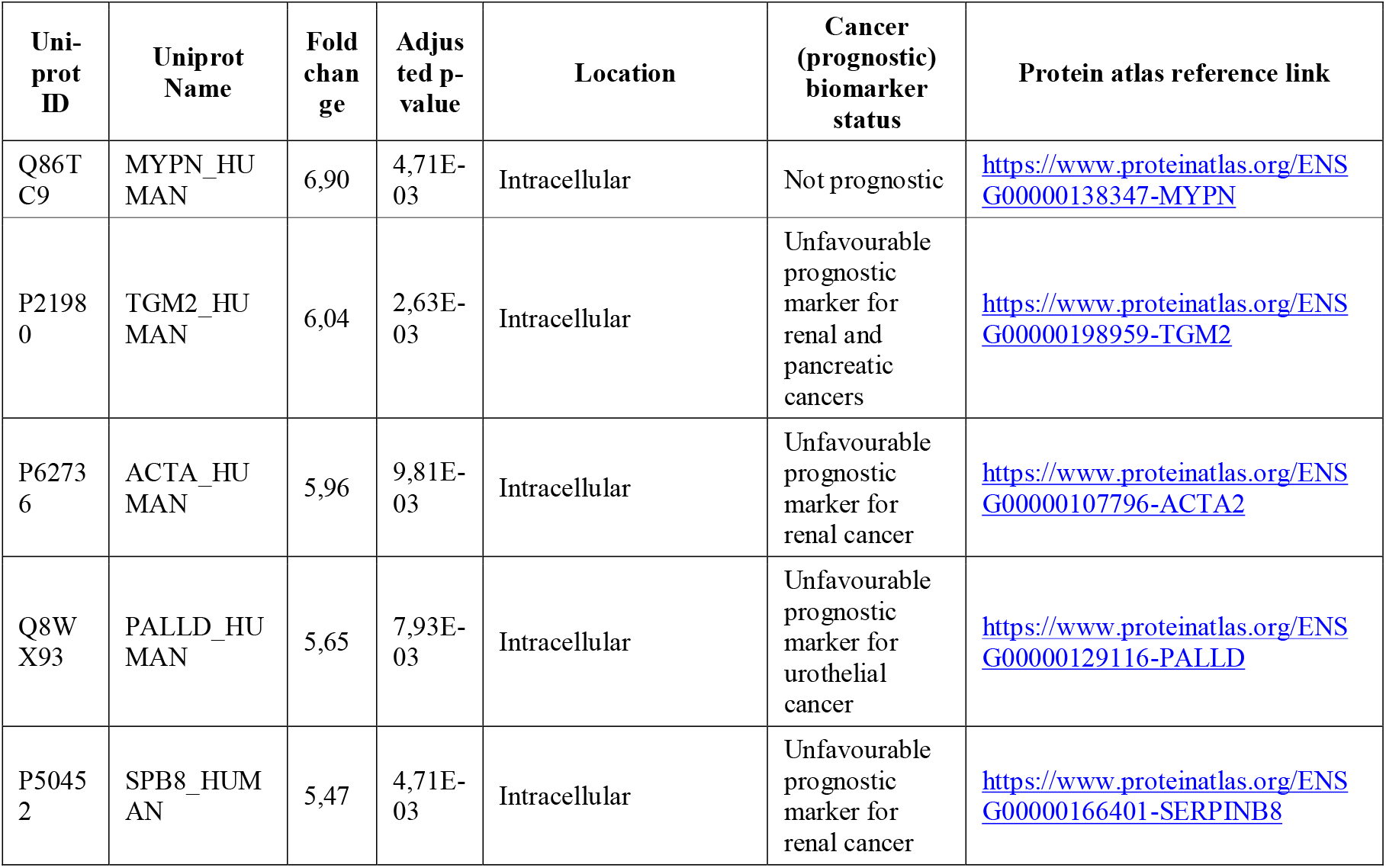
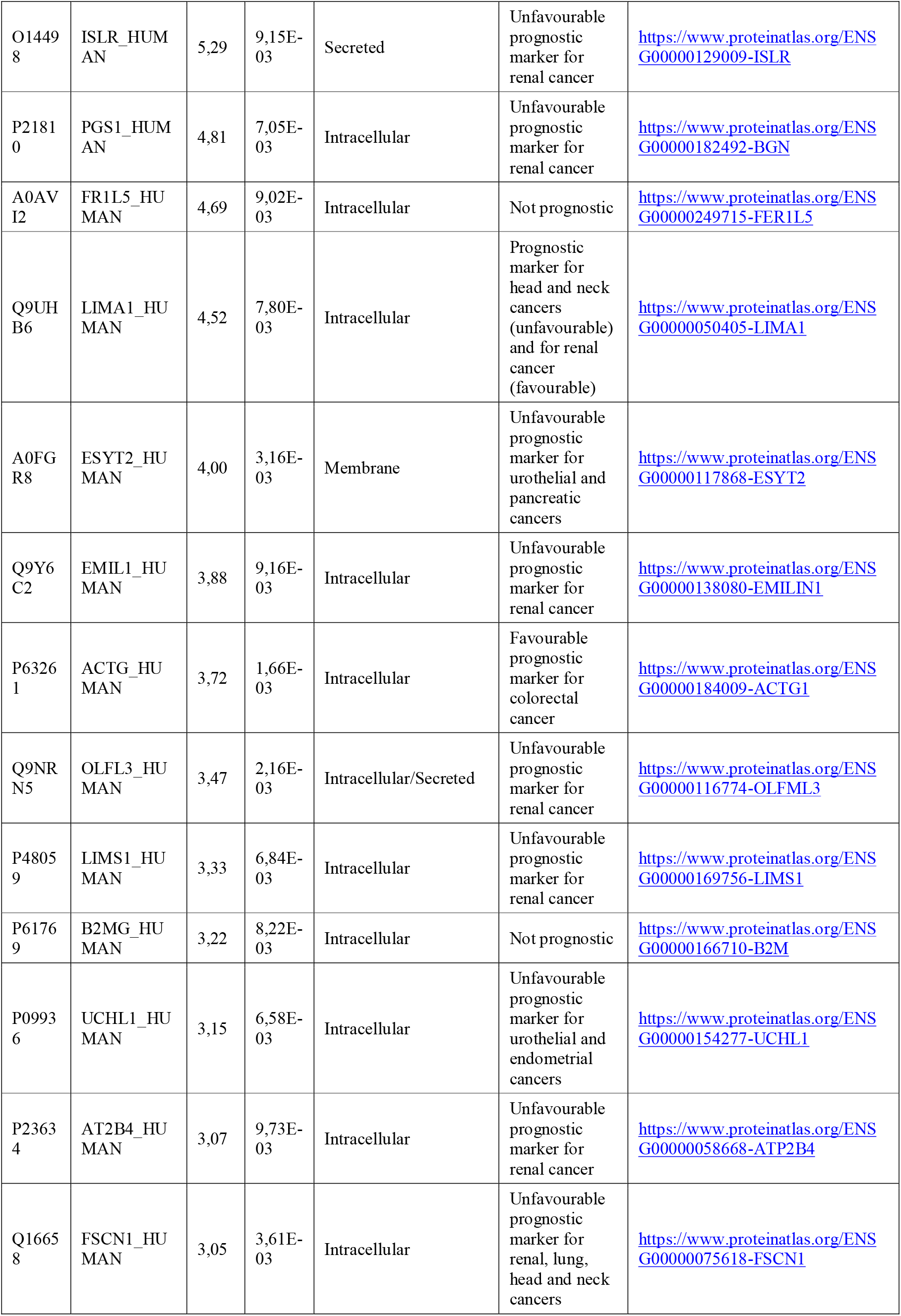
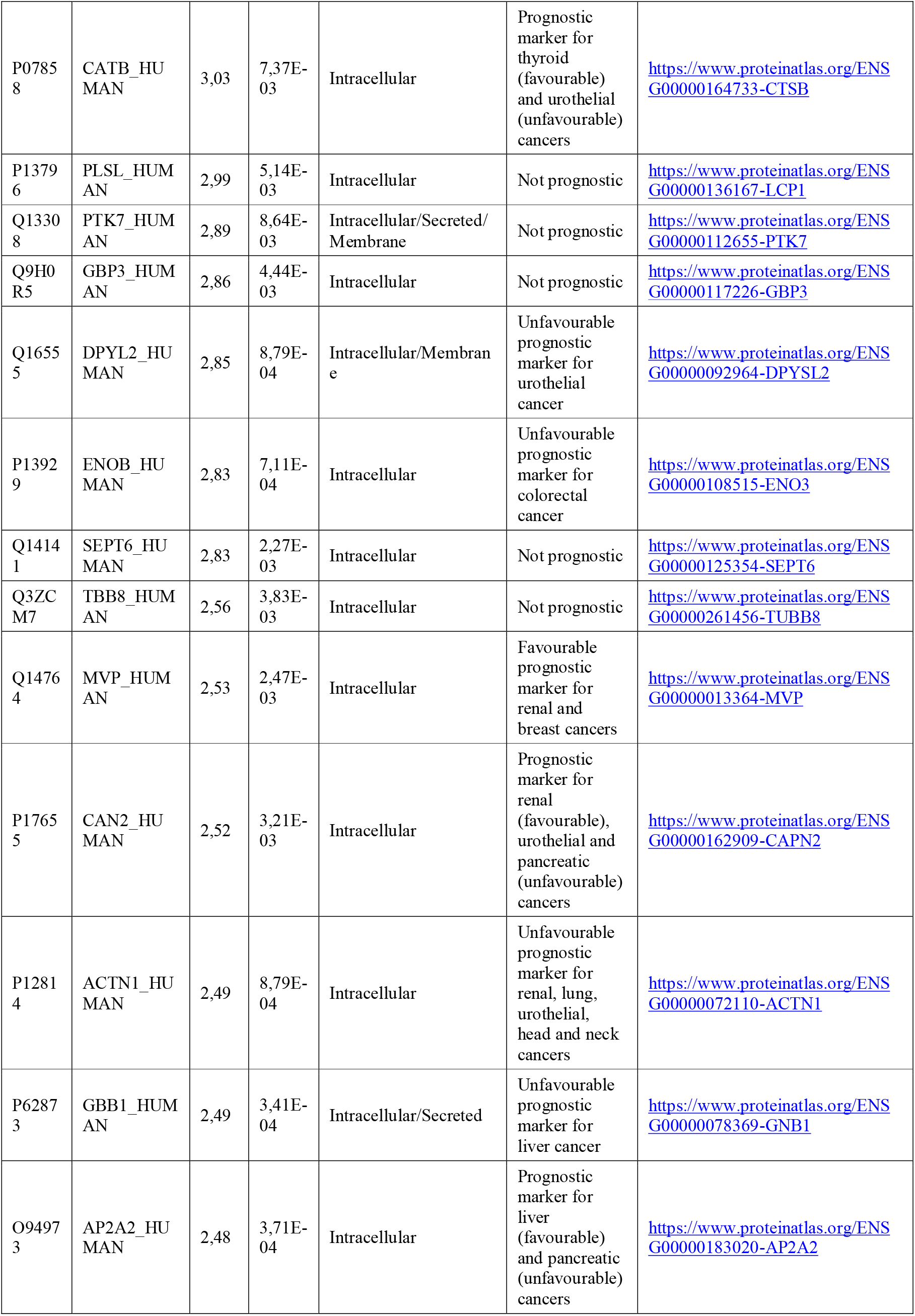
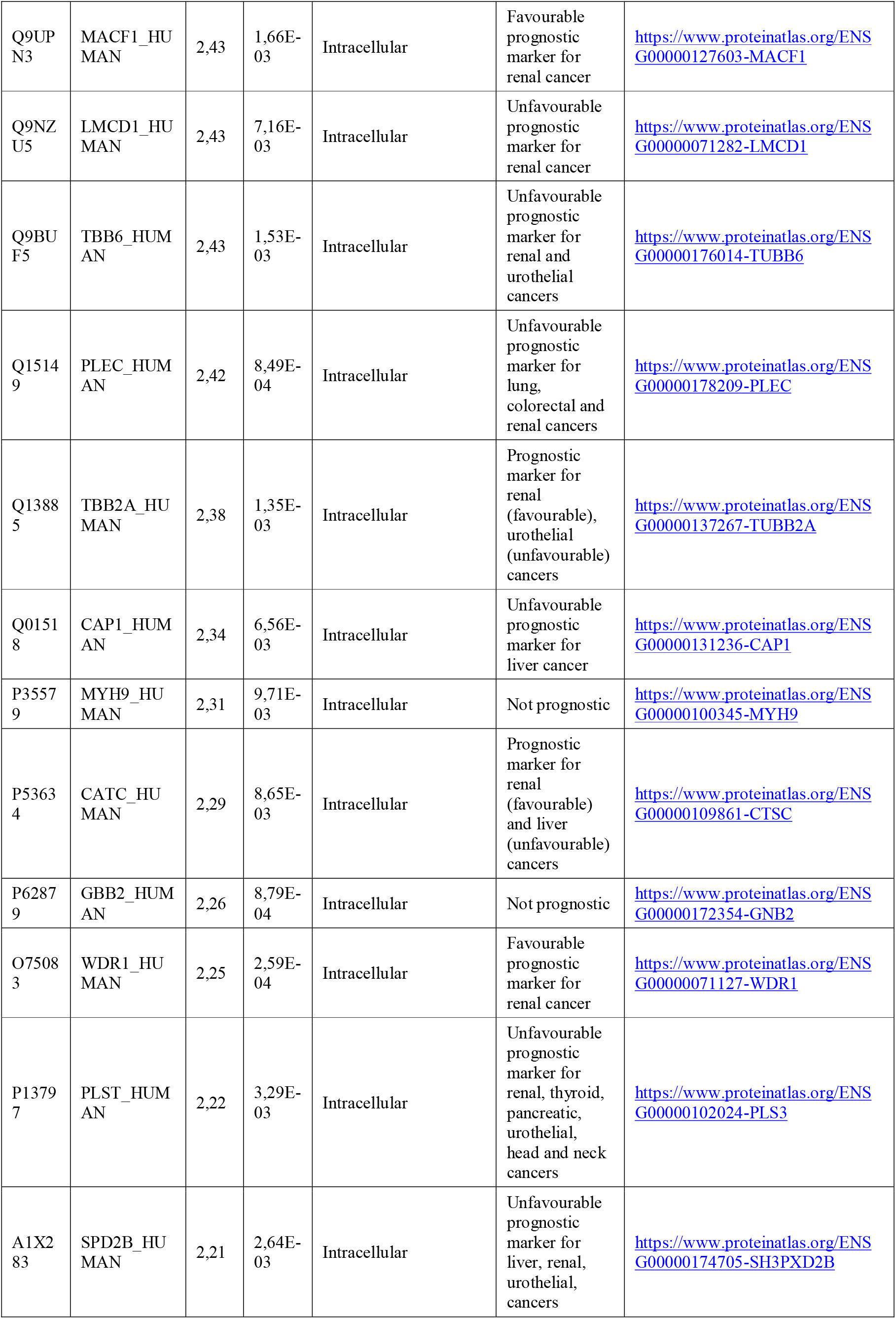
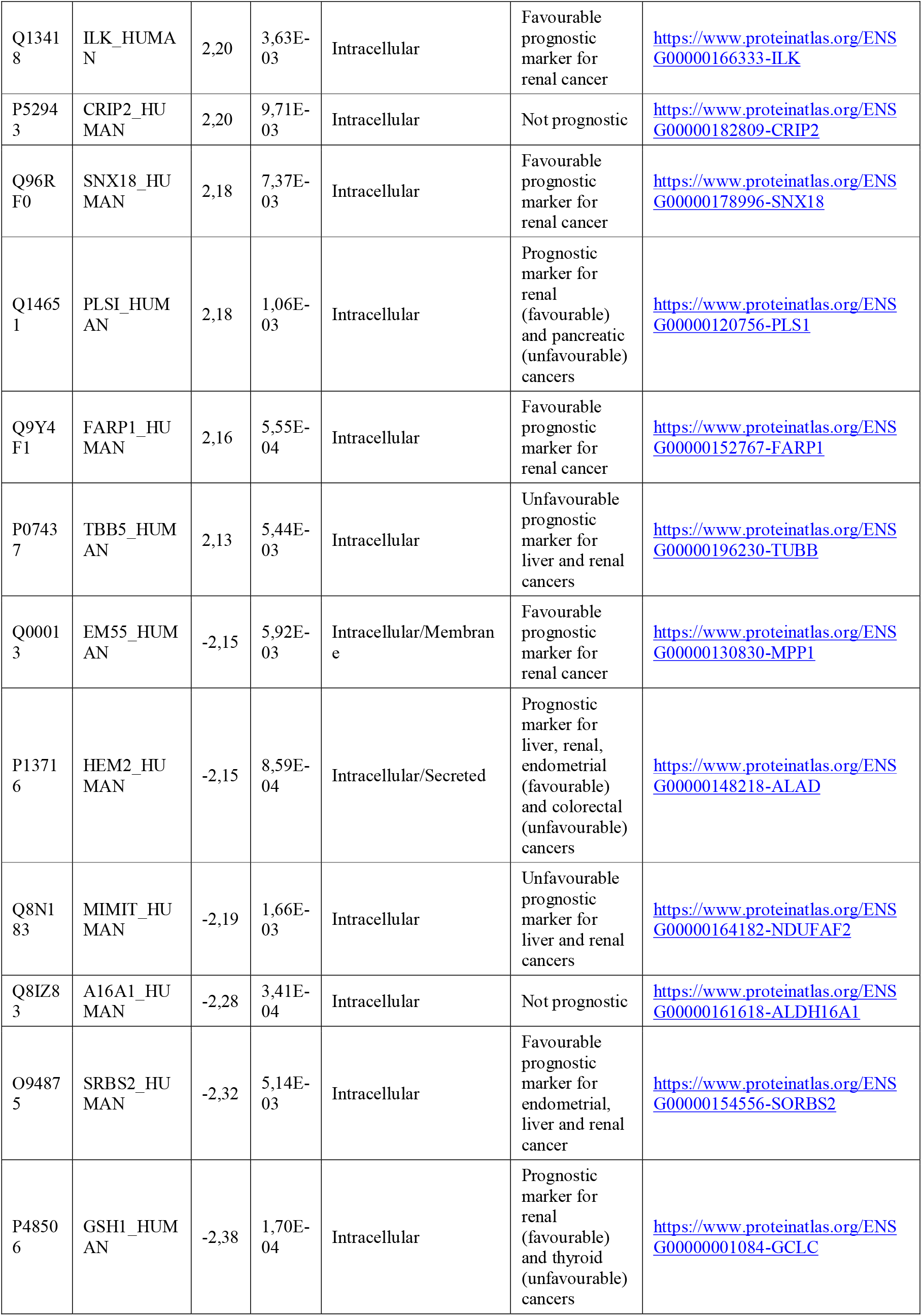
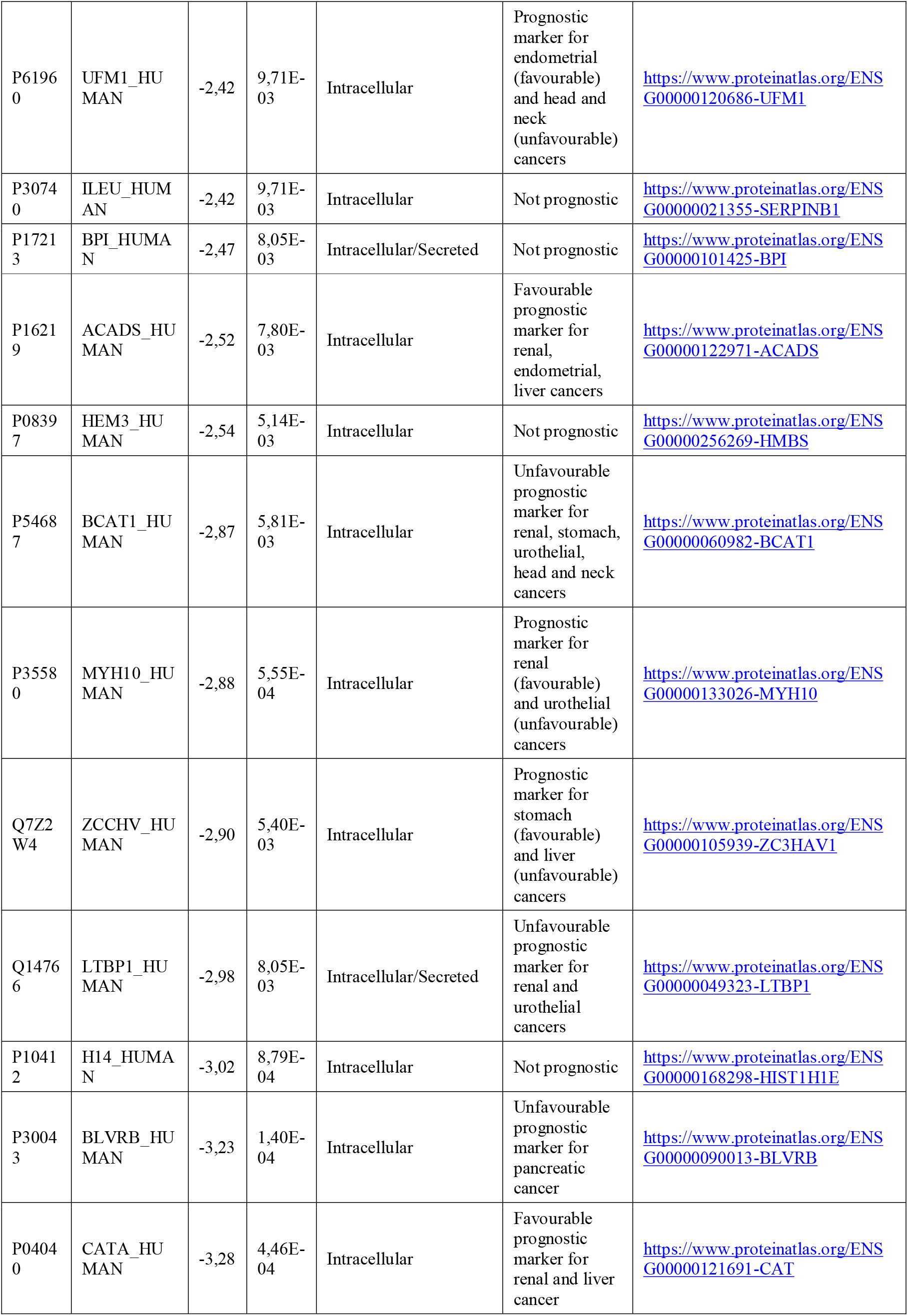
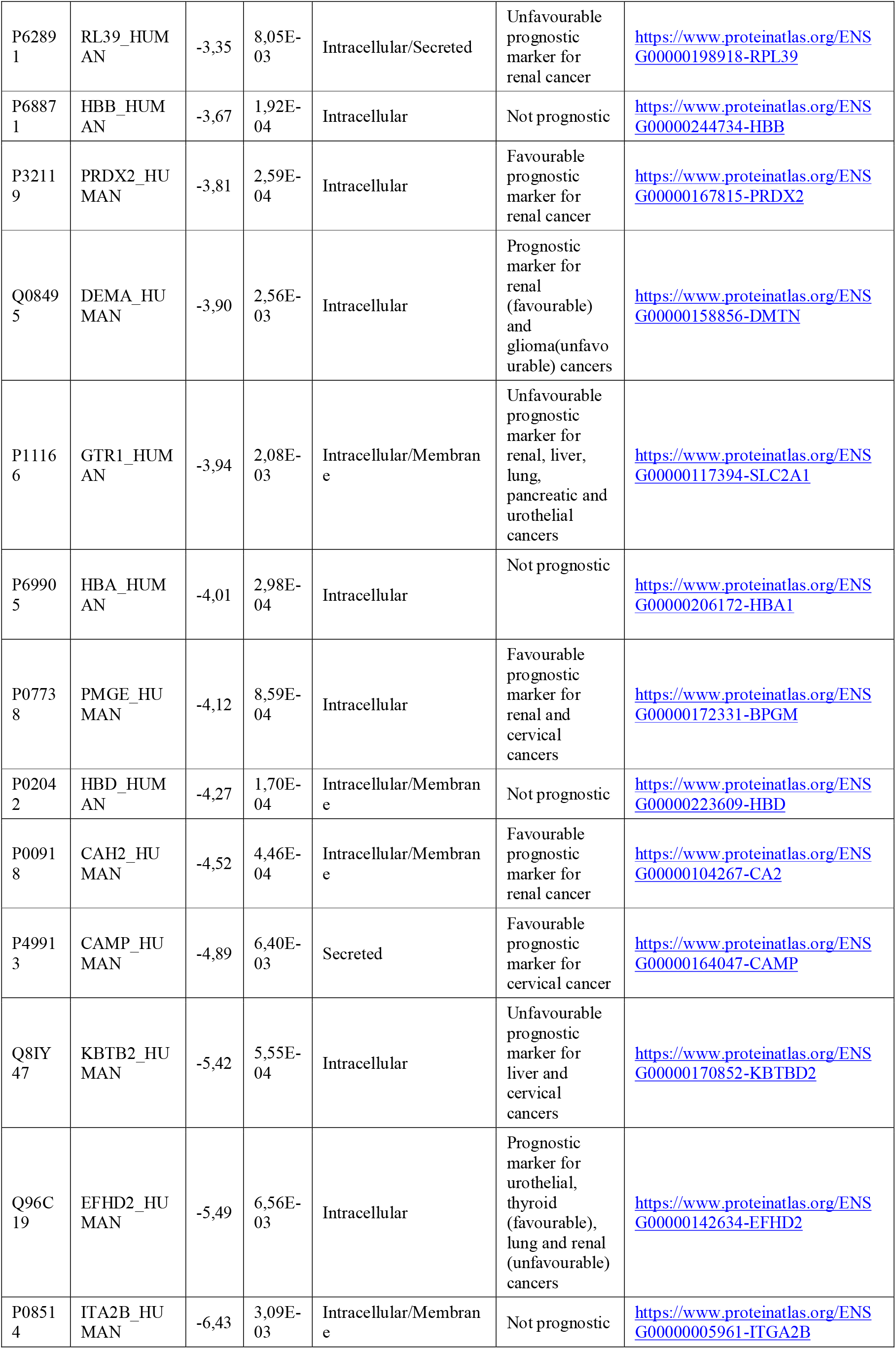
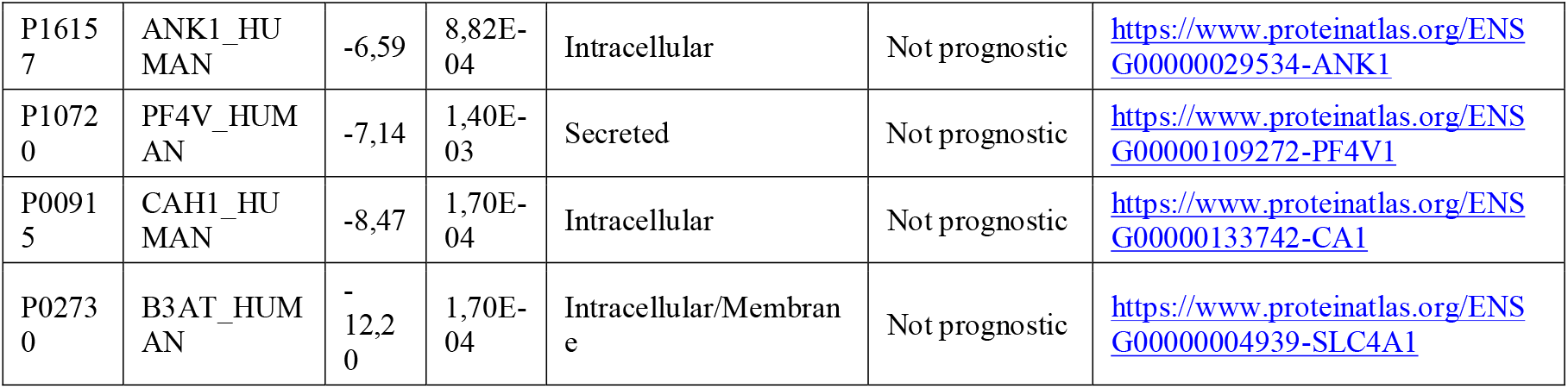
Differentially expressed proteins (fold change ≥2.1) measured in group comparison. Positive fold change describes proteins upregulated in the tumour group and negative fold change describes proteins downregulated in tumour compared to the normal adjacent group.

**Table S3:**
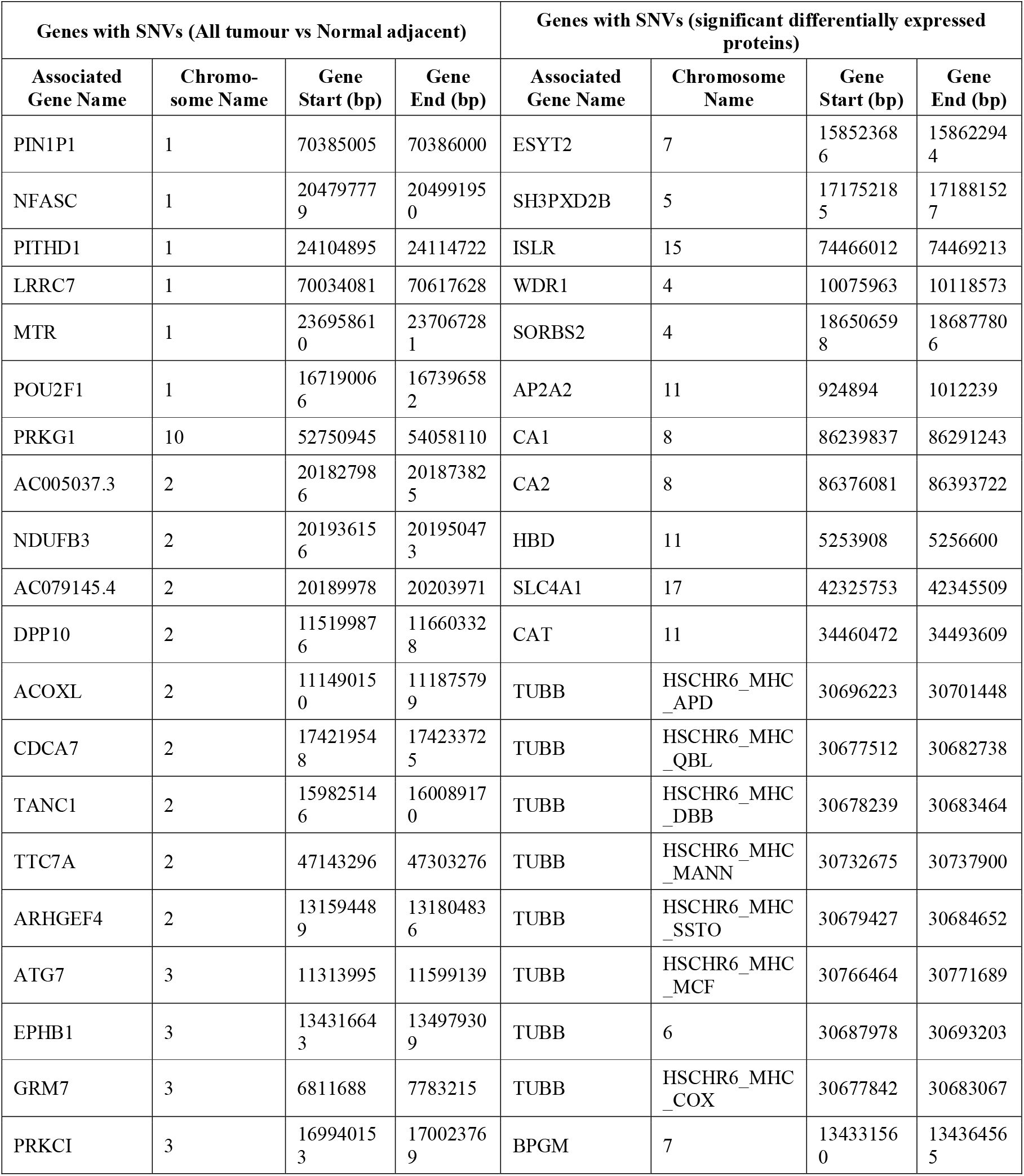
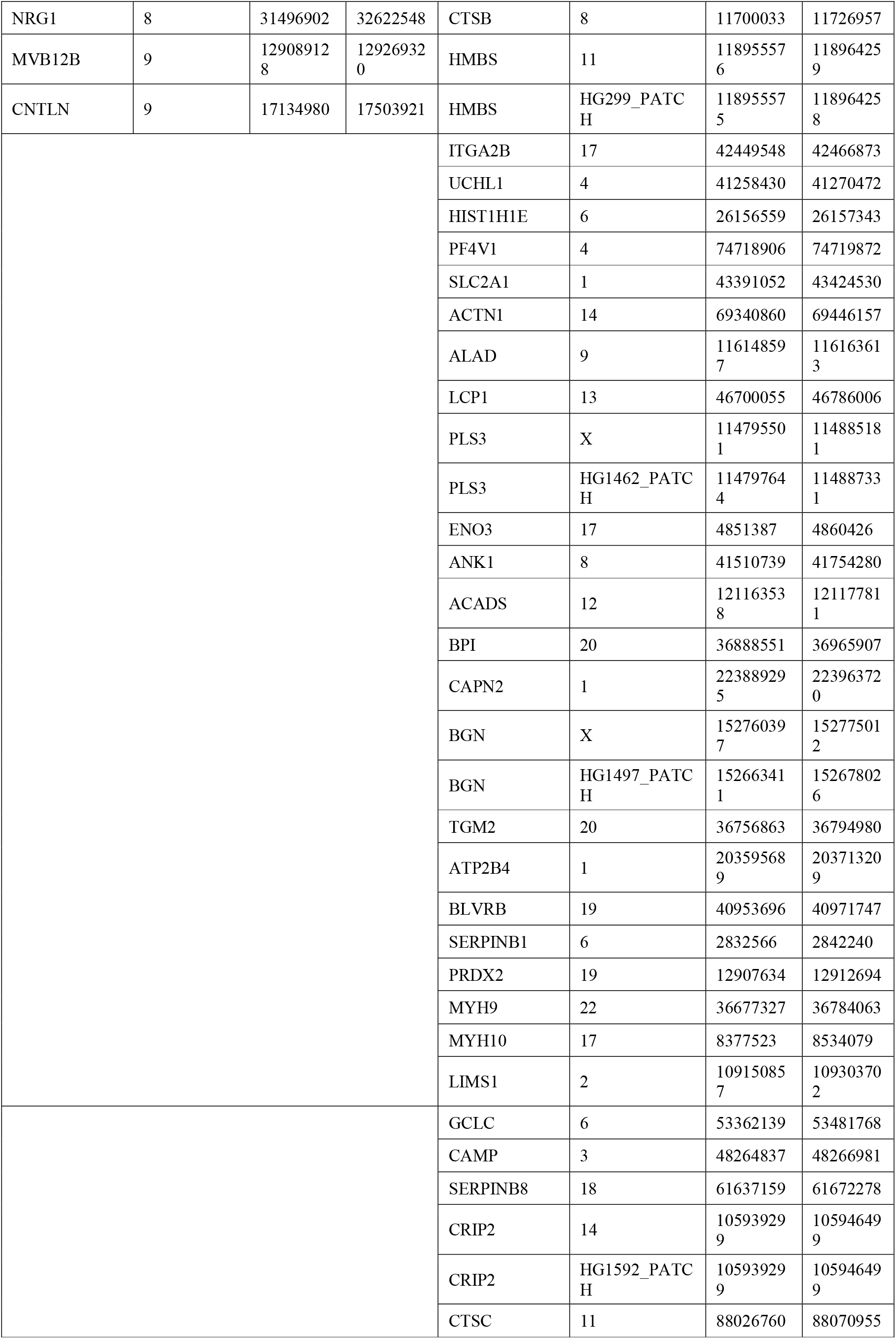
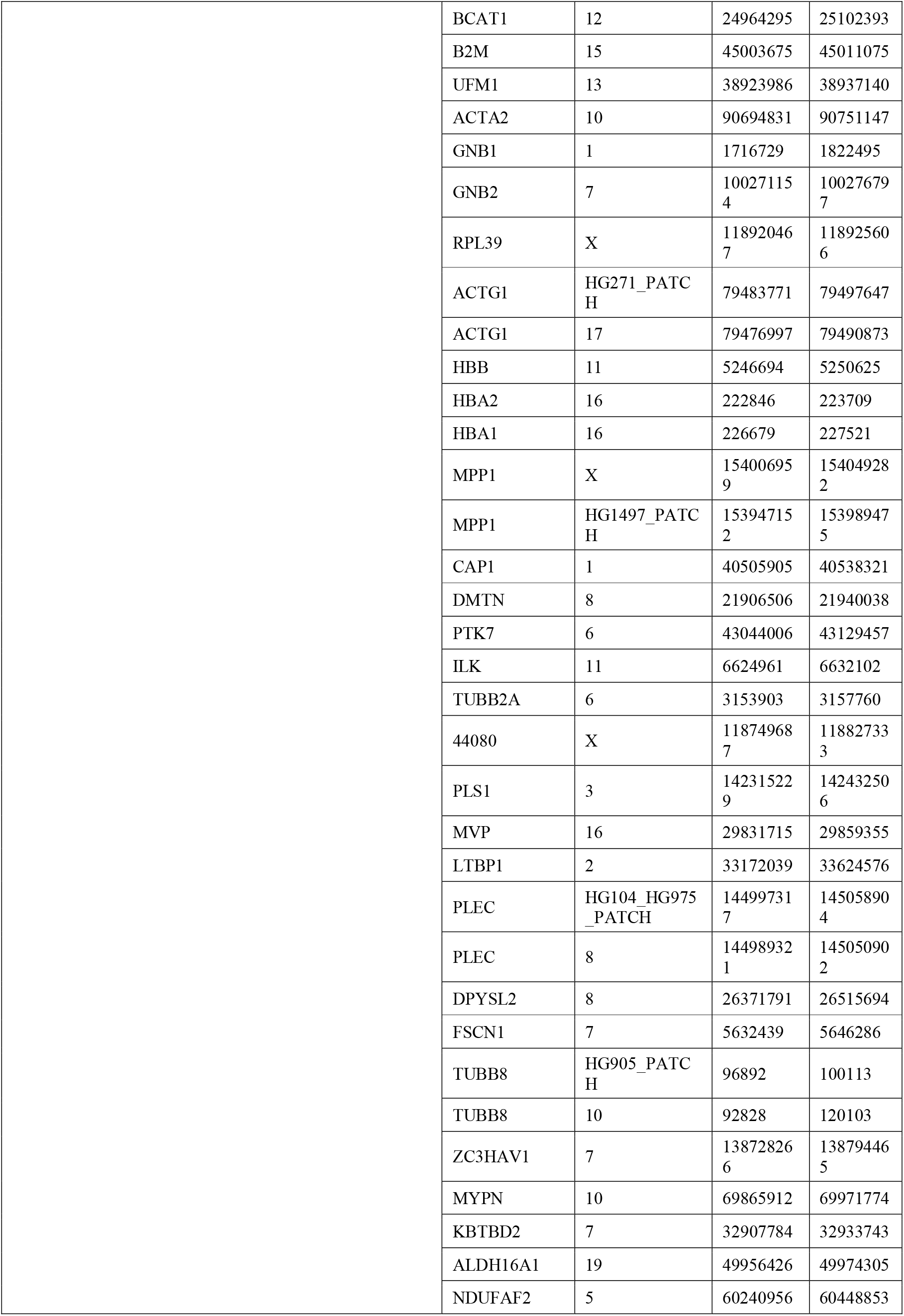
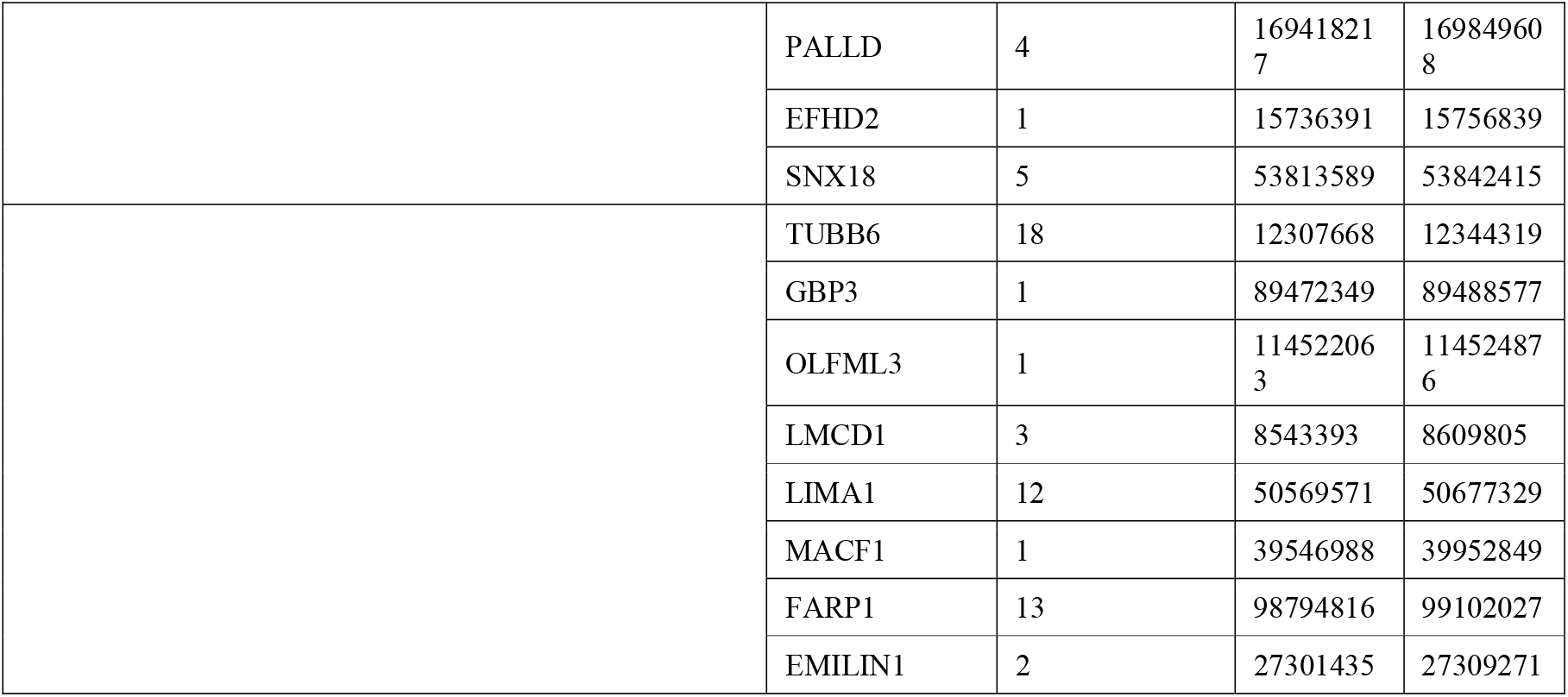
Top single nucleotide polymorphisms (SNVs) and associated genes

